# Quantitative Gait Categorization in Parkinson’s Disease with and without Freezing of Gait

**DOI:** 10.64898/2026.06.11.26355473

**Authors:** Suzann Donovan, Richa Tripathi, Hillary Chu, Doug Bernhard, Stewart A. Factor, J. Lucas Mckay, Christine D. Esper

**Affiliations:** Emory University College of Arts C Sciences, Neuroscience and Behavioral Biology Program; Jean and Paul Amos Parkinson’s Disease and Movement Disorders Program, Emory University School of Medicine, Department of Neurology; Emory University School of Medicine, Department of Biomedical Informatics

**Author notes:** Equal contributions (J. Lucas Mckay, Christine D. Esper).

**Keywords:** Parkinson’s disease, freezing of gait, motion analysis, gait analysis

## Abstract

**Background:** Freezing of gait (FOG) is a disabling and often underrecognized feature of Parkinson’s disease (PD). Objective gait analysis may improve characterization of this motor symptom.

**Objective:** To compare quantitative 3D gait parameters in PD with FOG (PD_F_) and PD without FOG (PD_NF_) in a routine clinical cohort. Methods: We retrospectively analyzed a sequential sample of 180 patients with PD referred for motion analysis between 2020 and 2024. All patients underwent 3D motion capture in the off-medication state. Eighteen gait outcomes spanning pace, rhythm, postural control, variability, and asymmetry domains were derived from steady-state walking tasks. FOG status was determined using physician documentation and Movement Disorder Society Unified Parkinson’s Disease Rating Scale (MDS-UPDRS) items. Group differences between PD_F_ (n=99) and PD_NF_ (n=81) were evaluated using independent samples t-tests, with outcomes adjusted for disease duration and corrected for multiple comparisons. A secondary analysis among PD_F_ compared those in Hoehn and Yahr (HCY) stage ≥III to those in HCY ≤II. Results: PD_F_ had longer disease duration, higher OFF MDS-UPDRS III scores, and higher Hoehn and Yahr stage than PD_NF_ but were similar in age and sex. After adjusting for disease duration and multiplicity, PD_F_ demonstrated reduced step length, stride length, and forward velocity, and greater cadence variability, while most postural control, and asymmetry measures were comparable between groups. Among PD_F_, advanced HCY stage was associated with impaired pace and rhythm, similar to previous reports among PD in general. Conclusion: In this large, sequential, clinically referred cohort, FOG was associated with more advanced PD and specific impairments in pace and gait variability. These findings support comprehensive 3D gait analysis as an objective tool to better delineate FOG-related gait abnormalities and identify features that may predict FOG, informing targeted interventions.

## Introduction

Parkinson’s disease (PD) is the second most common neurodegenerative disorder^1^ with motor features including bradykinesia, rest tremor, rigidity, postural instability, and gait abnormalities.^2^ Freezing of gait (FOG) is a particular gait disorder in PD that is defined as paroxysmal episodes resulting in an inability to step effectively, despite attempting to do so.^3^ Affecting up to 60% of PD patients, FOG increases the risk of falls and associated morbidity, contributing to reduced functional mobility, social isolation, and diminished quality of life.^4,5^ Its unpredictable nature makes clinical detection challenging,^6^ and current treatment options remain inadequate.

FOG and postural instability often co-exist although this is not a consistent finding. It is clear that they can influence each other behaviorally, however the exact nature of this relationship remains largely unknown.^7^ Attempts to address this association have included the study of postural control in patients with and without FOG; those with FOG been shown to have reduced postural control by several measures compared to PD patient without FOG and healthy controls. (Schlenstedt et al, Bekksarra et al). Furthermore, it has been suggested that postural instability interferes with gait symmetry such that it may actually lead to the development of levodopa unresponsive FOG and falls.^8^ Finally, there have been attempts to compare FOG with postural instability finding that that they have different demographic, non-motor and genetic predictors indicating that they are pathophysiologically distinct.^9^ To our knowledge, there have been no studies explicitly comparing gait outcomes in FOG with and without postural instability.

A providers’ ability to assess FOG in the clinical setting is limited, as episodes may not occur during routine visits.^6^ The Movement Disorder Society Unified Parkinson’s Disease Rating Scale (MDS-UPDRS)^10^ is the standard clinical scale for PD, but only 2 out of 65 items address FOG: a patient/caregiver question (2.13) and a clinician-rated observation (3.11), each scored 0-4 to reflect severity. While useful, these tools can miss transient or subjective manifestations such as FOG.^11^ FOG can also be assessed using the New Freezing of Gait Questionnaire, although this self-report instrument has acknowledged limitations, particularly potential recall bias and limited sensitivity to transient or context-dependent freezing episodes.^12^ Given these challenges, there is a need to better characterize PD patients with FOG using objective gait analyses, which may reveal measurable general gait abnormalities in PD Freezers. Accordingly, a range of objective tools have been used to assess FOG in primarily research settings, including video analysis,^13^ pressure-sensitive mats,^14^ inertial sensors,^15,16^ or 3D kinematics.^17–19^

The objective of our study is to compare quantitative 3D gait kinematics of PD patients with FOG (PD_F_) vs. PD patients without FOG (PD_NF_) in a large sample of consecutive patients recruited from our center. Our hypothesis is that PD_F_ will have reduced step length, stride length, and gait velocity. Because the sample size allowed further evaluation within the FOG group, an exploratory secondary objective was to repeat the analysis among patients with FOG to compare those with postural instability (Hoehn and Yahr^20^ stage III or higher) with those without postural instability (Hoehn and Yahr stage II or lower).

## Materials & Methods

### Data sources

PD patients are routinely seen in our Emory Brain Health Center Motion Analysis Laboratory (MAL) as part of their clinical assessments for potential neuromodulation interventions. We evaluated gait outcomes in all patients with PD (ICD-10 G20) whose diagnoses were confirmed by manual review of electronic health records adjudicated by trained abstractors, and who were seen in the MAL between February 2020 and July 2024. All patients were assessed in the practically defined off-medication state (i.e. off medications ≥ 12 hours).^21^ Patient motion analysis (MA) procedures were selected sequentially during this period. This cross-sectional retrospective study was approved by Emory University School of Medicine Institutional Review Board as STUDY00002688.

### 3D motion capture and data processing

All patients were assessed in the MAL which has a 150 m^3^ capture area (**Figure 1**) using a Motion Analysis Cortex 3D motion analysis system (software version 7.2.6.1828) with a standard 60 marker set described previously.^17,22,23^ Patients were assessed using standard protocols for assessing tremor and gait. During gait assessment, participants completed a minimum of five laps along a 14-foot walkway at their self-selected pace. Following completion of the walking laps, turning was assessed as a separate task, with participants performing turns within designated boxes, which were recorded and analyzed independently.

**Figure 1.**
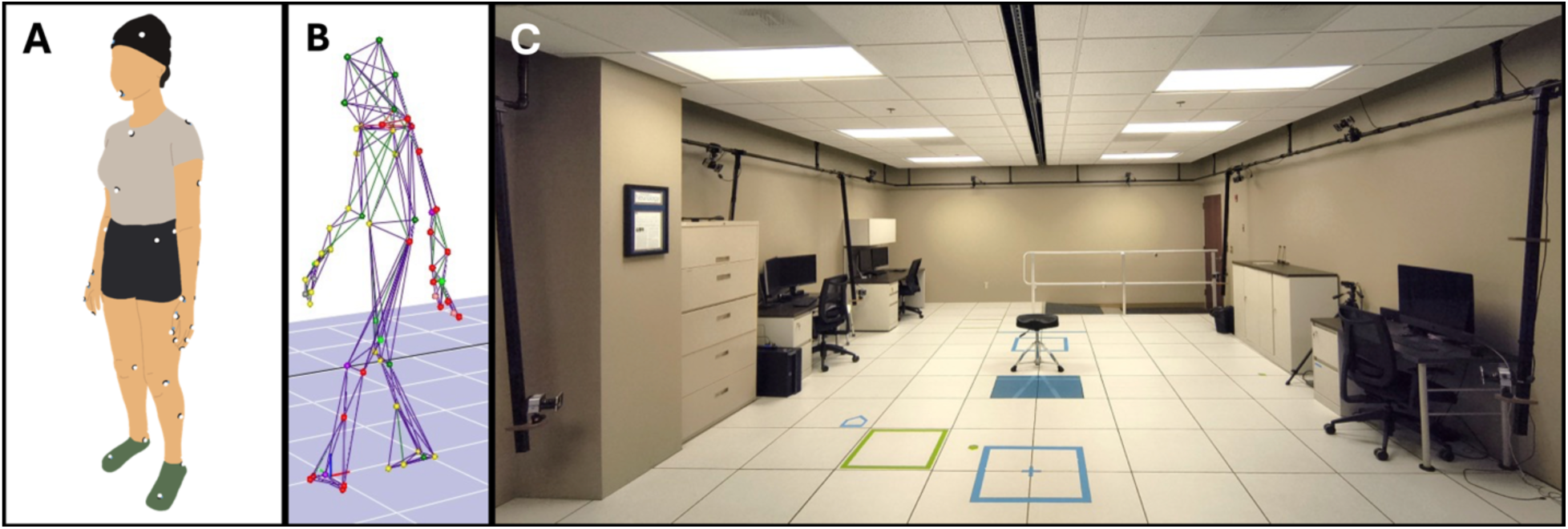
Clinical motion capture facility. Our center uses a custom set of 60 retroreflective kinematic markers for most cases (A). After data collection, commercial software triangulates the 3-dimensional coordinates of each kinematic marker in the laboratory space and assembles a deidentified wire frame or representation of the individual, preserving privacy (B). Our center measures ∼650 square feet and is used for both clinical and research applications (C).

Marker trajectories were sampled at 120 Hz; joint angles, segment positions, and temporal-spatial parameters were computed in Orthotrak according to manufacturer algorithms, with gait events (initial contact, toe-off) identified automatically from kinematic data and reviewed for accuracy by lab staff. Complete gait cycles were extracted and averaged to generate representative measures for each participant, which were summarized in tabular NRM files. Scalar outcomes included step length (cm), stride length (cm), forward velocity (cm/s), cadence (steps/min), total support time (%), swing phase (%), initial double support time (%), single support time (%), and step width (cm). For each variable and participant, average values, standard deviations, and asymmetry indices 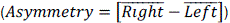 were computed. Outcomes were then organized into the domains of pace, rhythm, postural control, variability, and asymmetry. For parsimony, 18 of 28 potential outcomes were selected *a priori* for analysis (**Table S1**).

### FOG status

PD patients in the study were classified as either with FOG (PD_F_) or without FOG (PD_NF_). The following criteria were selected to best characterize FOG at the time of the MA procedure. Patients were classified as PD_F_ if they met any of the following criteria:

1. FOG documented in MA procedure note by a movement disorders trained physician (RT, CDE)
2. FOG identified in movement disorders clinic visit notes at any time within one year prior to MA visit
3. MDS-UPDRS 2.13 “freezing” score greater than 0 in movement disorders clinic visit notes at any time within one year prior to MA visit
4. MDS-UPDRS 3.11 “freezing of gait” score greater than 0 in movement disorders clinic visit notes at any time within one year prior to MA visit

PD_NF_ were identified as patients that did not meet any of these criteria.

### Analytic plan

Differences in clinical and demographic variables between PD_F_ and PD_NF_ were assessed with univariate tests (t-tests, chi-squared) as appropriate. For the primary analysis, group mean values across gait outcomes were initially visualized using radar plots, generated in R with the *ggradar* package after scaling each variable to a 0–100% range. For all other analyses, the original unscaled values were retained. Group differences were subsequently evaluated using independent-samples t-tests for each of the 18 selected gait outcomes. To account for imbalances in disease progression between PD_F_ and PD_NF_, each outcome was residualized from a linear regression on disease duration, and t-tests were repeated on the residuals. Multiple comparisons were controlled using the Bonferroni–Holm procedure. Summary statistics are reported as mean±SD. All analyses were conducted in R with α=0.05. Finally, to inform power calculations for future studies, differences in outcomes between groups were expressed as Cohen’s d,^24^ defined as

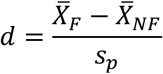

Where 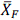 and 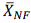 are the group means for PD_F_ and PD_NF_, respectively, and *s*_*p*_ is the pooled standard deviation. The pooled standard deviation combines the variability of both groups weighted by sample size.

For the exploratory secondary analysis, all primary analysis procedures were repeated within the FOG subgroup to compare participants with postural instability (Hoehn and Yahr stage ≥III) versus those without postural instability (Hoehn and Yahr stage ≤II).

## Results

### Patient population

Initial abstraction identified data of N=278 unique patients. Records were excluded from analyses for the following reasons: status-post deep brain stimulation (DBS) or focused ultrasound (FUS) (N=64); additional underlying gait impact other than FOG that could affect measurements, such concurrent spine or musculoskeletal issues, (N=30); gait testing not performed due to safety concerns (N=4). After exclusions, N=180 records were available for analysis. A flowchart depicting patient flow through the study is shown in **Figure 2**.

**Figure 2.**
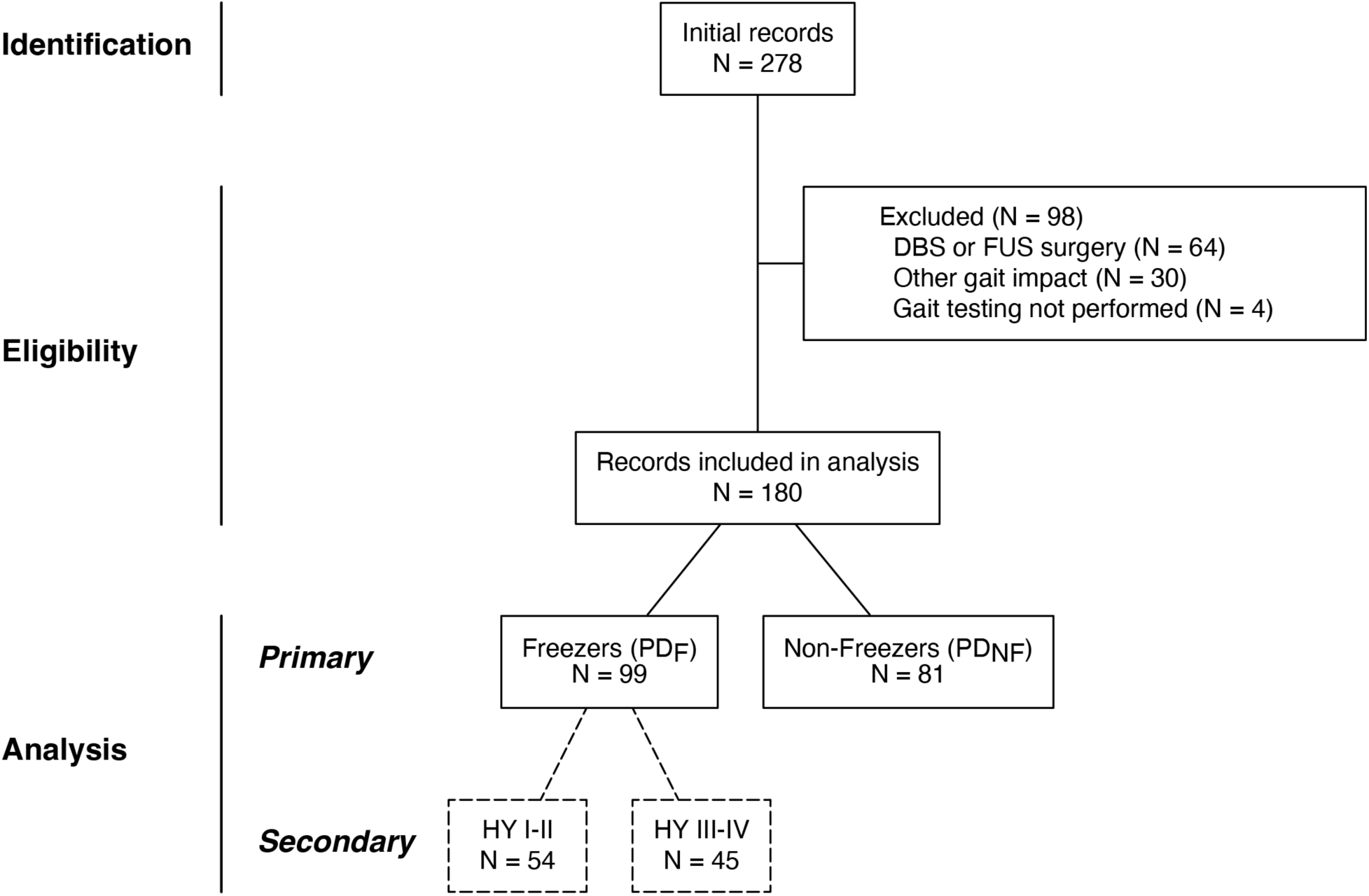
Flowchart summarizing how patient records were selected and analyzed in the study.

Clinical and demographic characteristics for the primary analysis are shown in **Table 1**. PD_F_ and PD_NF_ groups were similar in age (P = 0.21) and sex (P = 0.17), but PD_F_ had longer disease duration (P < 0.01), higher MDS-UPDRS-III scores (P < 0.01), and more advanced Hoehn and Yahr (HCY) stage (P < 0.01). Clinical and demographic characteristics for the secondary analysis are shown in **Table S2**.

**Table 1.** Clinical and demographic characteristics of the study sample, stratified by FOG status.

|  | PD<sub>F</sub> | PD<sub>NF</sub> | P value |
| --- | --- | --- | --- |
| N | 99 | 81 |  |
| Age |  |  | 0.21 |
| Mean (SD) | 64.4 (9.9) | 66.4 (10.8) |  |
| Range | 37.1 - 87.1 | 34.1 - 82.8 |  |
| Sex |  |  | 0.17 |
| Female | 31 (31%) | 18 (22%) |  |
| Male | 68 (69%) | 63 (78%) |  |
| Disease Duration |  |  | < 0.01 |
| Mean (SD) | 11.5 (4.7) | 9.3 (5.5) |  |
| Range | 2.1 - 27.4 | 1.4 - 29.9 |  |
| MDS-UPDRS-III |  |  | < 0.01 |
| Mean (SD) | 46.3 (12.6) | 40.0 (12.0) |  |
| Range | 20 - 75 | 18 - 64 |  |
| Hoehn and Yahr Stage |  |  | < 0.01 |
| I | 0 (0%) | 2 (2%) |  |
| II | 54 (55%) | 69 (85%) |  |
| III | 39 (39%) | 10 (12%) |  |
| IV | 6 (6%) | 0 (0%) |  |

Abbreviations: PD<sub>F</sub>, PD with FOG; PD<sub>NF</sub>, PD without FOG; MDS-UPDRS-III, Movement Disorder Society – Unified Parkinson’s Disease Rating Scale, Part III (Motor Examination). Clinical and demographic characteristics of patients included in the secondary exploratory analysis are summarized in **Table S2**.

Among PD_F_, patients in HCY stage ≥III were similar in age (3.5 years older on average, P = 0.08), sex (38% vs. 26% female, P = 0.21), and disease duration (12.3 vs. 11.1 years, P = 0.21) to those in HCY stage ≤II but had higher MDS-UPDRS-III scores (52.9 vs. 40.8 points; P < 0.01).

### Gait data

The total gait dataset included 180 separate files yielding 18 separate gait outcomes each, for a total of 3,240 outcomes derived from records of N=180 separate patients. There was an average of 11.0±2.9 (range, 4-24) observations (steps or strides) for each gait outcome. All available outcomes were included in analyses.

### Primary Analysis: comparison of gait outcomes between PD_F_ and PD_NF_

Crude comparisons revealed statistically significant impairments in 9 of 18 gait outcomes in the Pace, Rhythm, Variability, and Asymmetry domains (**Table 2**). After adjusting for disease duration and applying Bonferroni–Holm correction, significant differences remained in Pace (reduced Step Length, P<<0.001; Stride Length, P<<0.001; and Forward Velocity, <0.001;) and Variability (increased Cadence SD, P<0.01). Visualization with radar plots (**Figure 3**) confirmed this trend, with the largest differences observed in the Pace domain. In general, we found little qualitative change between the contrast curves observed before and after correcting for overall severity (**Figure 3**, compare dark Blue and Green vs. light Blue and Green lines). Effect sizes ranged from negligible (Stride Length Asymmetry, Cohen’s *d*=0.001) to medium (Step Length, *d*=0.671, Stride Length, *d*=0.672) according to cutoff values proposed by Cohen.^24^

**Figure 3.**
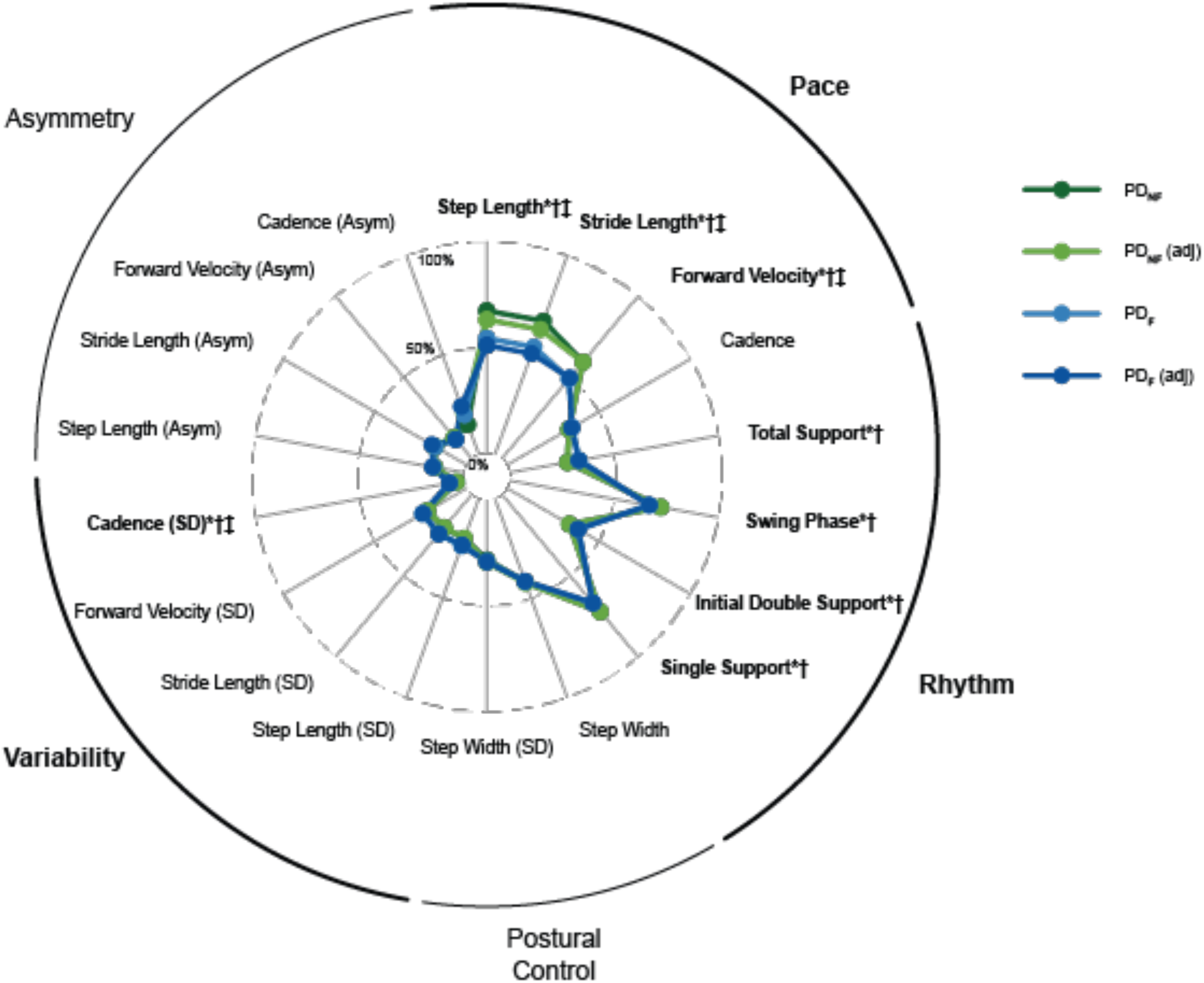
Radar plot visualizing contrasts between PD_F_ and PD_NF_ on gait outcomes. PD_F_ (adj) and PD_NF_ (adj) designate group averages for PD_F_ and PD_NF_ after removing effects of PD duration. Abbreviations: SD, standard deviation; Asym, asymmetry. *P<0.05, independent samples t-test. †P<0.05, independent samples t-test adjusted for disease duration. ‡P<0.05, independent samples t-test adjusted for disease duration and multiplicity of outcomes (Bonferroni-Holm procedure). Percentile lines (0, 50, 100) show the absolute range. Group means fall lower for variables influenced by extreme outliers (e.g., Cadence Asymmetry) and closer to 50% for more stable measures (e.g., Step Length).

**Figure 4.**
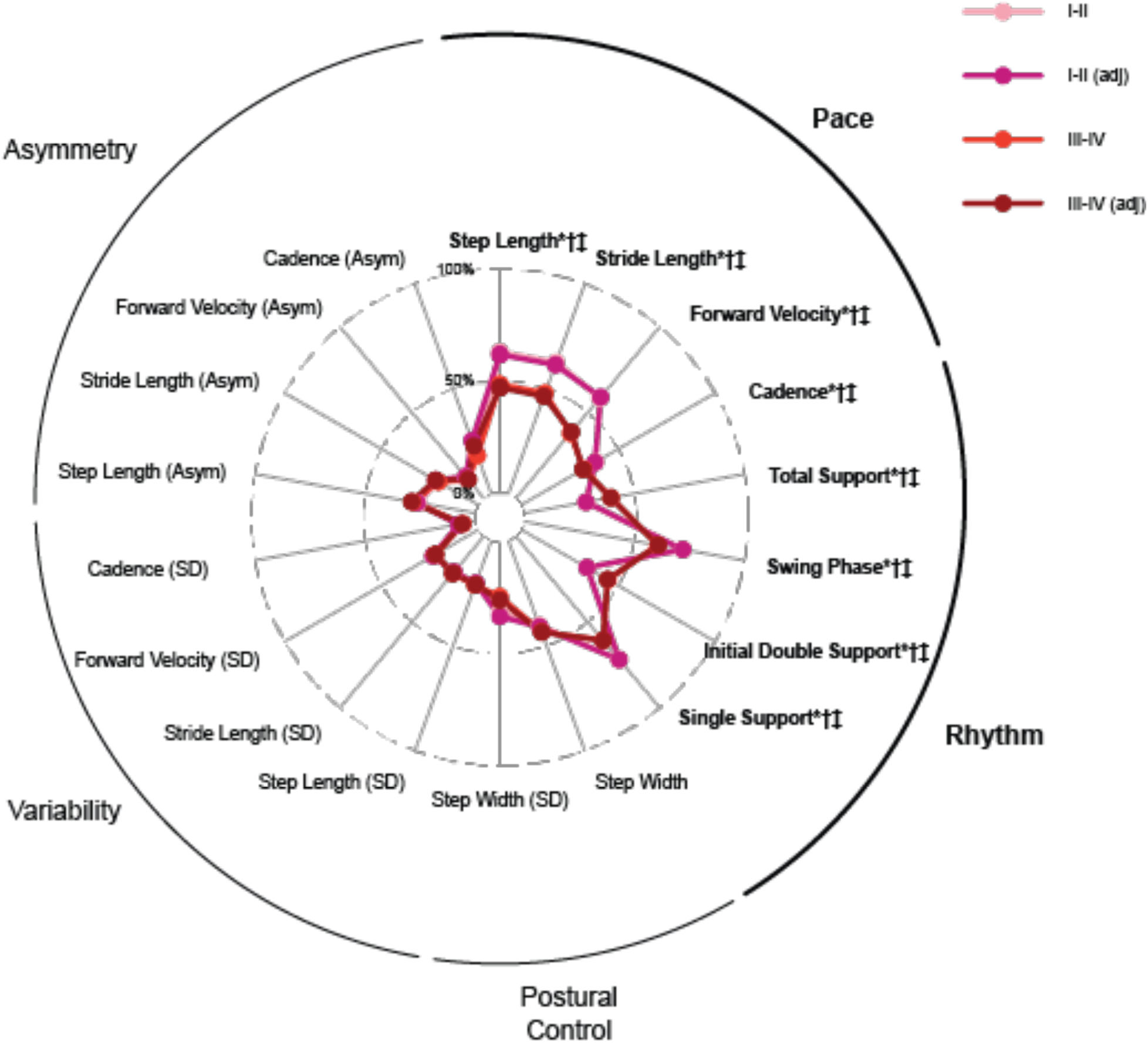
Radar plot visualizing contrasts between HCY ≥III and HCY ≤II on gait outcomes among PD_F_. (adj) designate group averages after removing effects of PD duration. Abbreviations: SD, standard deviation; Asym, asymmetry. *P<0.05, independent samples t-test. †P<0.05, independent samples t-test adjusted for disease duration. ‡P<0.05, independent samples t-test adjusted for disease duration and multiplicity of outcomes (Bonferroni-Holm procedure).

**Table 2.**
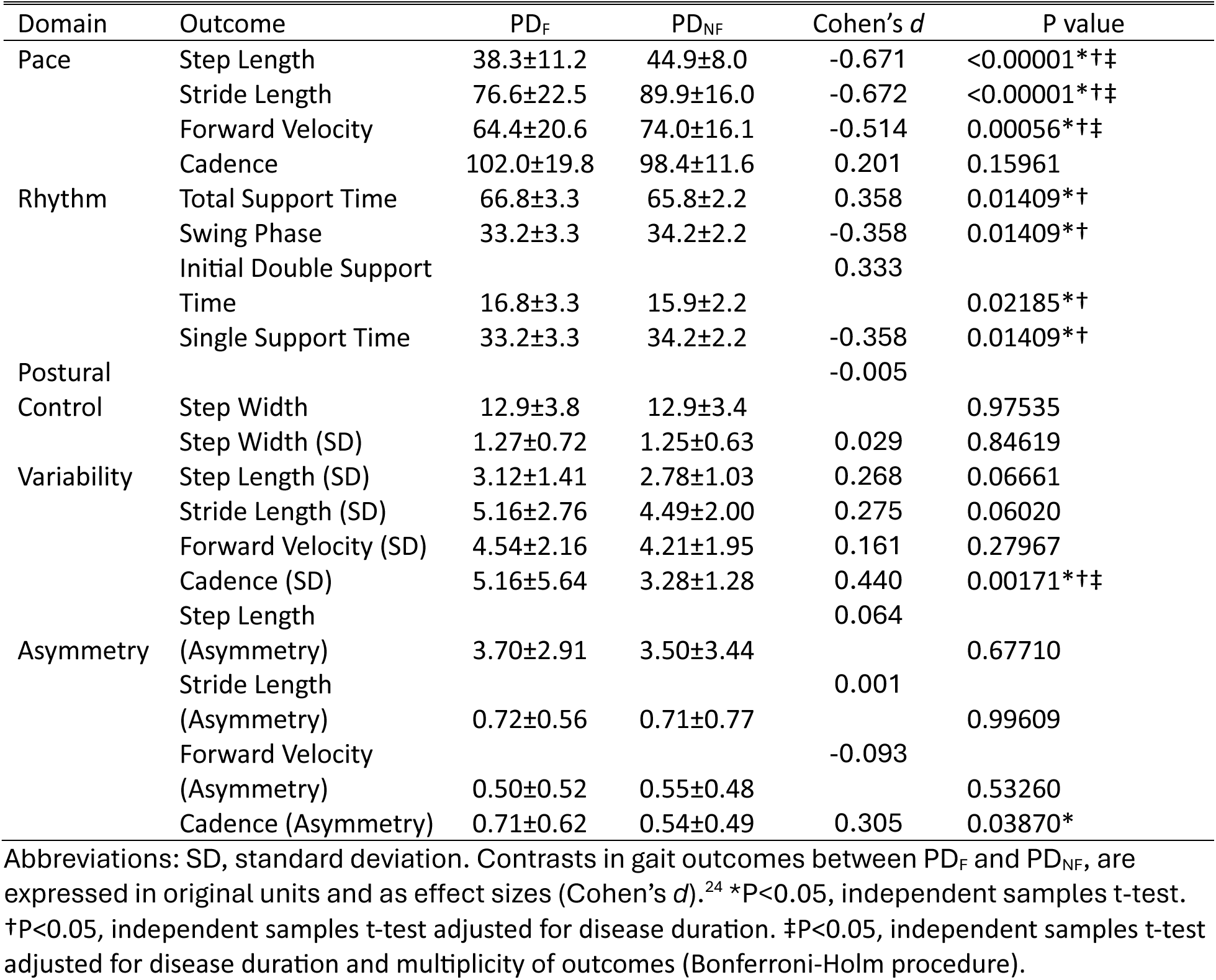
Contrasts in gait outcomes between PD_F_ and PD_NF_.

### Secondary Analysis: comparison of gait outcomes between H&Y ≥ III and H&Y ≤ II among PDF

Crude comparisons revealed statistically significant impairments in all gait outcomes in the Pace and Rhythm domains (8 of 18 total outcomes; **Table 3**). After adjusting for disease duration and applying Bonferroni–Holm correction, significant differences remained across all Pace outcomes, including reduced Step Length (P=0.00132), Stride Length (P=0.00121), Forward Velocity (P<0.00001), and Cadence (P=0.00444) in participants with HCY III–IV compared with HCY I–II. Significant Rhythm-domain differences also persisted, including increased Total Support Time/reduced Swing Phase (both P=0.00183) and Initial Double Support Time (P=0.00146). No statistically significant differences were observed in the Postural Control, Variability, or Asymmetry domains. Effect sizes ranged from negligible (Stride Length SD, Cohen’s *d*=0.048; Step Length SD, *d*=0.053) to large (Forward Velocity, *d*=-1.022), with most significant Pace and Rhythm outcomes showing medium effects.

**Table 3.** Contrasts in gait outcomes between among PD_F_ between HCY III-IV and HCY I-II.

| Domain | Outcome | III-IV | I-II | Cohen's *d* | P value |
| --- | --- | --- | --- | --- | --- |
| Pace | Step Length | 34.4±10.39 | 41.5±10.9 | -0.665 | 0.00132*†‡ |
|  | Stride Length | 68.8±20.89 | 83.1±21.86 | -0.670 | 0.00121*†‡ |
|  | Forward Velocity | 54.1±16.64 | 72.9±19.69 | -1.022 | <0.00001*†‡ |
|  | Cadence | 95.7±17.02 | 107±20.72 | -0.578 | 0.00444*†‡ |
| Rhythm | Total Support Time | 67.9±3.229 | 65.9±3.002 | 0.652 | 0.00183*†‡ |
|  | Swing Phase | 32.1±3.229 | 34.1±3.002 | -0.652 | 0.00183*†‡ |
|  | Initial Double Support Time | 18±3.176 | 15.9±3.103 | 0.664 | 0.00146*†‡ |
|  | Single Support Time | 32.1±3.229 | 34.1±3.002 | -0.652 | 0.00183*†‡ |
| Postural Control | Step Width | 13.2±4.235 | 12.6±3.312 | 0.155 | 0.45396 |
|  | Step Width (SD) | 1.15±0.6337 | 1.38±0.7691 | -0.325 | 0.10417 |
| Variability | Step Length (SD) | 3.16±1.262 | 3.08±1.531 | 0.053 | 0.78875 |
|  | Stride Length (SD) | 5.23±2.541 | 5.1±2.952 | 0.048 | 0.81169 |
|  | Forward Velocity (SD) | 4.47±2.195 | 4.61±2.153 | -0.063 | 0.75672 |
|  | Cadence (SD) | 4.73±2.295 | 5.52±7.354 | -0.139 | 0.45995 |
| Asymmetry | Step Length (Asymmetry) | 3.87±3.098 | 3.57±2.766 | 0.104 | 0.61304 |
|  | Stride Length (Asymmetry) | 0.706±0.522 | 0.727±0.5937 | -0.036 | 0.85525 |
|  | Forward Velocity (Asymmetry) | 0.46±0.3343 | 0.53±0.6354 | -0.135 | 0.48383 |
|  | Cadence (Asymmetry) | 0.67±0.6776 | 0.746±0.575 | -0.122 | 0.55328 |

Abbreviations: SD, standard deviation. Contrasts in gait outcomes between PD<sub>F</sub> and PD<sub>NF</sub>, are expressed in original units and as effect sizes (Cohen's *d*).<sup>24</sup> \*P<0.05, independent samples t-test. †P<0.05, independent samples t-test adjusted for disease duration. ‡P<0.05, independent samples t-test adjusted for disease duration and multiplicity of outcomes (Bonferroni-Holm procedure).

## Discussion

To our knowledge, this is the largest single-center study to date comparing quantitative gait parameters between PD_F_ and PD_NF_ groups, with a sample size of N = 180. By contrast, an informal review of comparable works^14,25–31^ yielded average sample sizes of 55±40 (range, 22 – 144), whereas the largest study of which we are aware (N = 310)^32^ did not consider FOG as a variable. This study demonstrated that PD_F_ have more advanced motor disease and exhibit characteristic gait abnormalities, including shorter step and stride length, slower forward velocity, and increased cadence variability than PD_NF_, even after accounting for disease duration, whereas age, sex, and several rhythm, postural control, and asymmetry measures did not differ _NF_, which was consistent with other FOG studies in the literature.^25,27,33–36^ Additionally, our study reported that PD_F_ vs. PD_NF_ were similar in age and sex.

Among FOG studies that reported demographic data, participants with FOG generally had a longer disease duration and greater motor disease severity, as reflected by the MDS-UDPRS III scores; however, HCY stage, another marker of disease severity, is often reported inconsistently and is rarely stratified by FOG status. In our study, HCY stage was included and demonstrated that PD_F_ had a more advanced HCY stage (p<0.01). Some studies have shown that patients with similar disease duration may be classified at very different HCY stages, and that disease duration alone is a weaker predictor of disability, progression, and survival as compared to clinical staging and/or motor scores.^37–39^ In addition, a tremor-predominant PD patient may have a higher MDS-UPDRS III score due to substantial tremor burden, yet exhibit minimal postural instability and therefore a lower HCY stage. A validation study of the HCY scale in over 3,000 patients found that tremor had the lowest correlation with HCY of any cardinal motor sign,^40^ underscoring the importance of reporting *both* the MDS-UPDRS III and HCY stage to fully characterize a patient’s motor status. HCY stage is also consistently used as a marker of gait, balance impairment, and fall risk in PD, with several studies demonstrating the importance of these outcomes.^41–43^ Overall, our findings align with prior studies and indicate that FOG’s association with longer disease duration and more severe PD underscores the importance of clearly categorizing this gait disorder and of future research on targeted interventions.

Among the FOG literature, step length and stride length emerge as one of the most consistently impaired gait parameters across multiple studies,^25,27,34,35,44,45^ similar to our findings. Gait velocity is also consistently reduced in PD_F_ vs. PD_NF_,^25,27,33,34,46^ which we also identified. Several prospective and observational studies report that shorter step/stride length and slower gait speed are key markers of fall risk in PD.^47–51^ Monitoring these parameters over the course of PD is important to predict future fall risks and pre-emptively intervene earlier when feasible.

While our study did not find statistically significant differences in cadence between PD_F_ versus PD_NF_ (roughly 4% higher in PD_F_, p<0.16), we did identify increased cadence variability (increased SD) was significant (roughly 20% higher in PD_F_, p<0.01). In the FOG literature, findings on cadence were more variable across studies. One study found no significant differences between PD_F_ vs. PD_NF_ in both the off and on medication states,^25^ while another study reported that PD_F_ had a greater decrease in cadence during turning compared to PD_NF_.^33^ In the context of turning, a meta-analysis found PD_F_ had significantly higher cadence with a mean difference of 10.1 steps/minute (95% CI: 4.87-15.33).^52^ This apparent contradiction may reflect task-specific differences, with PD_F_ showing increased cadence during challenging turning tasks but reduced cadence during prolonged turning.^33^ Earlier identification and targeted treatment of such higher risk maneuvers may reduce fall risk in PD patients.

Because FOG emerges later in the disease course in many people, it was somewhat surprising that PD_F_ were not older than PD_NF_ in this consecutively recruited cohort. In fact, if anything, PD_F_ were slightly *younger* than PD_NF_ (64±10 vs. 66±11 years; p=0.21). Previous FOG studies that included both groups and reported demographic characteristics have likewise found little or no age difference between participants with and without FOG.^25,33,53,54^ However, many of those studies were designed to recruit age-matched groups, making it difficult to determine whether the observed age similarity reflected the underlying clinical population or the sampling strategy. Our findings suggest that, at least in some consecutively recruited PD cohorts, the common impression that FOG is primarily a feature of older patients may be incomplete; FOG may also occur in relatively younger patients, and age differences by FOG status may be modest even without deliberate matching. This pattern may also reflect clinic-based sampling: patients experiencing the functional burden of FOG may present to specialty care earlier than expected, producing unanticipated age distributions even in consecutively recruited cohorts.

Existing literature on sex differences in FOG prevalence in PD remains limited and mixed, making the sex distribution observed in our sample of particular interest. The demographics of this consecutively recruited sample provided no evidence of an association between sex and FOG status. Specifically, the sex distribution did not differ significantly between the PD_F_ and PD_NF_ groups, although the PD_F_ group included a slightly higher proportion of females (31% vs. 22%; p = 0.17). This difference was not statistically significant and may be attributable to chance. Although the current literature remains limited with respect to sex distribution by FOG status in PD, prior reports have yielded mixed findings regarding sex-based differences in FOG prevalence, with some reporting no evidence of sex-based differences^55^ and others reporting evidence consistent with such differences.^56^ Notably, the possibility of a higher prevalence of FOG among women is particularly intriguing given that PD itself is generally less prevalent in women than in men, a pattern often attributed in part to the neuroprotective effects of estrogen on dopaminergic neurons.^57^ If replicated, this contrast could provide indirect support for the hypothesis that FOG may depend, at least in part, on nondopaminergic mechanisms. It is important to note that women with PD are underrepresented in clinical trials overall,^58–60^ moreover, they are particularly underrepresented in FOG intervention trials as compared to men.^55^ Therefore, when recruiting patients for such studies, deliberate efforts to enroll and retain women are essential to improve the generalizability and equity of FOG research.

Our secondary analysis comparing gait outcomes between participants with HCY ≥III and HCY ≤II among PD_F_ supports prior large-cohort observations^32,61^ that advanced PD is associated with slower walking, shorter steps and strides, and greater time spent in double support, and extends these findings to individuals with FOG. In our freezer subgroup, HCY III–IV participants showed a similar advanced-disease gait signature, with prominent Pace impairments and Rhythm-domain changes indicating greater reliance on support time. The absence of corrected differences in Variability, Asymmetry, and Postural Control measures suggests that, among individuals with FOG, HCY stage primarily differentiated pace and temporal support characteristics during this assessment, rather than producing global abnormalities across all gait domains. This pattern is consistent with the broader literature describing progressive PD gait impairment as involving reduced speed and step length, impaired rhythmicity, and increasing axial burden.

Although we were initially surprised not to observe differences in the postural-control domain of gait between participants with and without pull-test-defined postural instability on MDS-UPDRS III, this dissociation is not necessarily unexpected. Postural control during gait depends heavily on anticipatory postural adjustments, a feed-forward and internally timed process that shifts the center of mass over the support limb before and during stepping, whereas MDS-UPDRS item 3.12 (pull/retropulsion test) primarily probes reactive postural responses—automatic, externally triggered corrective stepping after an unexpected perturbation. These processes are related but not interchangeable, and prior work has argued against treating gait, balance, and posture deficits in Parkinson’s disease as a single construct.^62^ Bloem et al. similarly showed that pull test performance correlated only moderately with dynamic posturography, suggesting that the retropulsion test should not be interpreted as a general proxy for postural instability.^63^ Jacobs and Horak provided further evidence for separability: gait and pull test scores contributed independent information when predicting fall frequency, and a model including one-leg stance and gait—but not the pull test—best classified fallers versus non-fallers. In that context, our null finding in the gait postural-control domain does not materially weaken the interpretation; if anything, it is consistent with the idea that reactive postural instability and gait-related balance control reflect partially distinct constructs. In further support of this interpretation, we also found that abnormality on pull-test-like balance tests was independent of the presence of FOG.^64^

Our study has several limitations. Because our patients were recruited from a routine clinical screening process for neuromodulation intervention (DBS and FUS), they predominantly had moderate-to-advanced PD, which may limit the generalizability of our findings across the full spectrum of disease severity. Another limitation of our study is that we only included steady state walking as part of our routine motion capture protocol, which prevents us from considering objective gait abnormalities during other more complex movements such as turning,^33^ systematic variation of turn angles,*^65^* gait initiation,*^CC^* dual-task paradigms,*^C7,C8^* and backward walking.^27,34^ In addition, our gait assessment was performed only in the off medication state. Although dopaminergic medications are known to influence gait – particularly by improving pace*^CS^* – it remains unclear whether the FOG-related differences observed here would persist when patients are assessed in the ON state.

Despite these limitations, this study has several strengths. At N=180, to our knowledge, this is the largest single center study evaluating differences in quantitative parameters between PD_F_ and PD_NF_ cohorts. A brief review of several comparable studies^14,25–27,27–30^ identified sample sizes ranging between 22 and 82. Gait kinematics were measured via 3D motion capture, which serves as the reference gold standard in laboratory settings.^34,52,70^ In addition to spatiotemporal gait parameters, 3D motion capture provides kinematic data, including 3D joint angles across the gait cycle at the pelvis, hip, knee, angle, and upper limb segments – information not available from modalities such as gait mats or standard video analysis. Although this was not the primary focus of our study, these additional variables may further distinguish PD_F_ from PD_NF_ and ultimately enhance the identification and treatment of FOG. Furthermore, our use of a sequential, clinically referred patient population reduces selection bias and enhances the generalizability of our findings to routine PD care. Lastly, our study analyzed 18 distinct gait outcomes, including measures from the variability and asymmetry domains, providing a more comprehensive assessment of gait analysis in FOG as compared to other prior studies with a narrower set of outcomes.

## Conclusion

In summary, this study demonstrated that PD patients with FOG have more advanced motor disease and characteristic gait abnormalities – shorter step and stride length, slower forward velocity, and greater cadence variability – even after adjusting for disease duration. Our use of gold-standard 3D motion capture, a large sequential clinical cohort, and a broad set of 18 gait outcomes across multiple domains provides a detailed and clinically relevant description of gait in PD with and without FOG. Our findings underscore the importance of incorporating objective gait measures alongside traditional clinical scales when evaluating patients with suspected FOG and when counseling patients about fall risk and mobility.

Future work should extend these findings in several directions. Longitudinal studies are needed to determine whether specific steady-state gait metrics, particularly pace and variability measures, can serve as early markers of imminent FOG and future falls, and how these metrics change over the course of PD. Additional studies should integrate joint angle data as well as more challenging gait tasks—such as turning, gait initiation, dual-task walking, and backward walking—into 3D motion capture and wearable-sensor protocols to capture task-specific gait signatures that may better reflect real-world FOG. Interventional trials are also warranted to test whether pharmacologic, neuromodulatory, and rehabilitative strategies differentially modify quantitative gait parameters in PD_F_ versus PD_NF_, and whether changes in these parameters translate into meaningful clinical benefit. Finally, future studies should prioritize adequate enrollment of women and other underrepresented groups and examine how sex, cognition, and comorbidities interact with quantitative gait measures and FOG severity to support more personalized approaches to treatment and prognosis in PD.

## Data Availability

All data produced in the present study are available upon reasonable request to the authors

## Supplemental Material

**Table S1.** Mapping of scalar gait outcomes from MotionAnalysis software to gait domains. A bullet (•) indicates inclusion of an outcome in a given domain in analyses. Outcomes without bullets were excluded from analyses for parsimony.

| Statistic | Outcome | Domain |  |  |  |  |
| --- | --- | --- | --- | --- | --- | --- |
|  |  | Pace | Rhythm | Postural Control | Variability | Asymmetry |
| Average | Step Length (cm) | • |  |  |  |  |
|  | Stride Length (cm) | • |  |  |  |  |
|  | Forward Velocity (cm/s) | • |  |  |  |  |
|  | Cadence (steps/min) | • |  |  |  |  |
|  | Total Support Time (%) |  | • |  |  |  |
|  | Swing Phase (%) |  | • |  |  |  |
|  | Initial Double Support Time (%) |  | • |  |  |  |
|  | Single Support Time (%) |  | • |  |  |  |
|  | Step Width (cm) |  |  | • |  |  |
| Standard Deviation | Step Length (cm) |  |  |  | • |  |
|  | Stride Length (cm) |  |  |  | • |  |
|  | Forward Velocity (cm/s) |  |  |  | • |  |
|  | Cadence (steps/min) |  |  |  | • |  |
|  | Total Support Time (%) |  |  |  |  |  |
|  | Swing Phase (%) |  |  |  |  |  |
|  | Initial Double Support Time (%) |  |  |  |  |  |
|  | Single Support Time (%) |  |  |  |  |  |
|  | Step Width (cm) |  |  | • |  |  |
| Asymmetry | Step Length (cm) |  |  |  |  | • |
|  | Stride Length (cm) |  |  |  |  | • |
|  | Forward Velocity (cm/s) |  |  |  |  | • |
|  | Cadence (steps/min) |  |  |  |  | • |
|  | Total Support Time (%) |  |  |  |  |  |
|  | Swing Phase (%) |  |  |  |  |  |
|  | Initial Double Support Time (%) |  |  |  |  |  |
|  | Single Support Time (%) |  |  |  |  |  |
|  | Step Width (cm) |  |  |  |  |  |

**Table S2.**
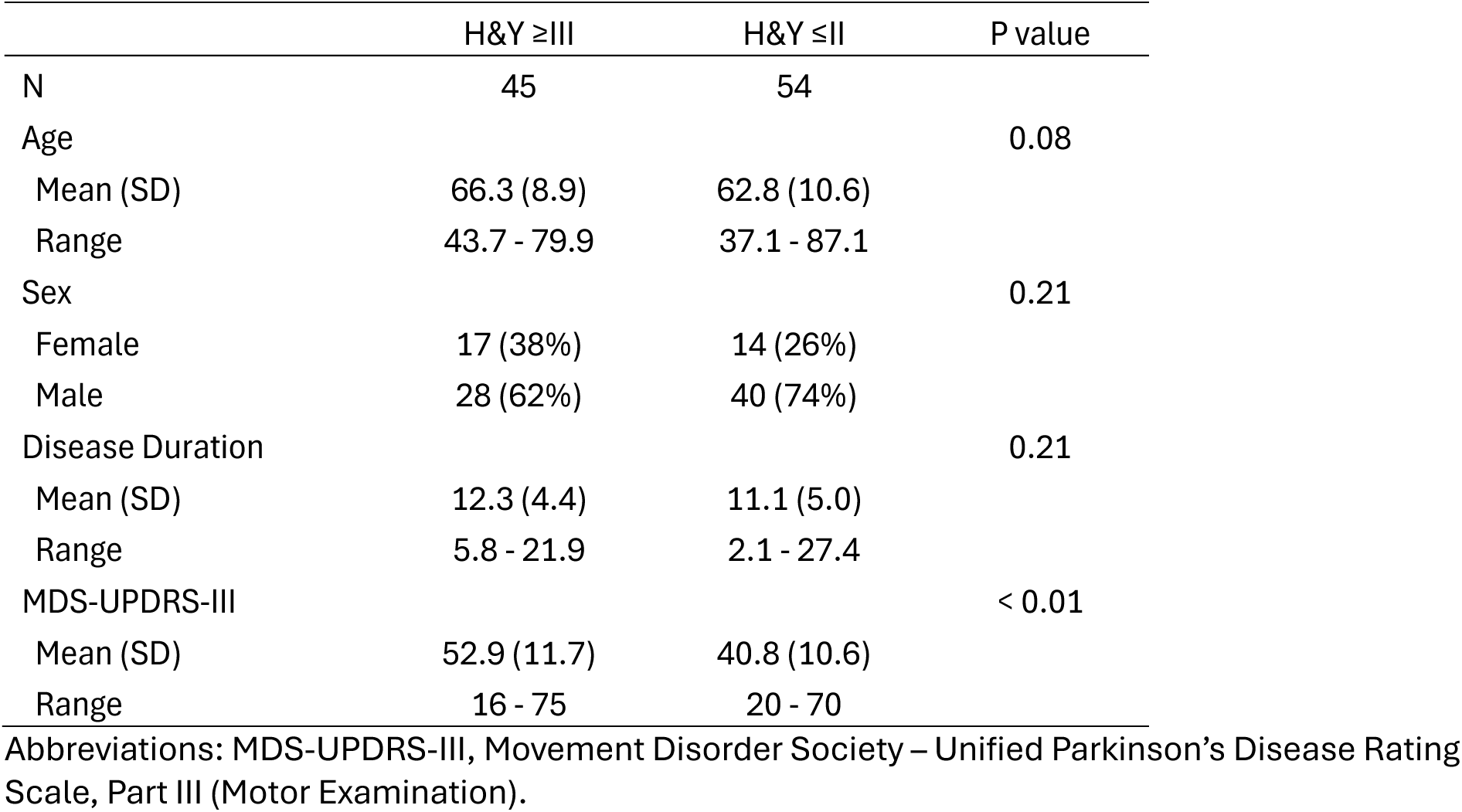
Clinical and demographic characteristics of PDF patients in the study sample, stratified by presence of postural instability.

## Notes

### Competing Interest Statement

The authors have declared no competing interest.

### Author Declarations

IRB of Emory University gave ethical approval for this work

### Funding

- The McCamish Parkinson’s Disease Innovation Program at Georgia Tech

